# Humoral and cell-mediated response in colostrum after exposure to severe acute respiratory syndrome coronavirus 2

**DOI:** 10.1101/2021.01.03.20248715

**Authors:** Vignesh Narayanaswamy, Brian Pentecost, Dominique Alfandari, Emily Chin, Kathleen Minor, Alyssa Kastrinakis, Tanya Lieberman, Kathleen F. Arcaro, Heidi Leftwich

**Author notes:** California State Assembly, Committee on Education, Capitol Office, Sacramento, CA 94249, USA. Indicates co-senior author (Kathleen F. Arcaro, Heidi Leftwich).

## Abstract

**Background:** Colostrum provides an immune sharing between a mother and her infant. The transfer in colostrum of antibodies against SARS-CoV-2 and the elicited cytokines may provide crucial protection to the infant. There is limited literature on the immune response to SARS-CoV-2 present in colostrum.

**Objective:** To evaluate the presence of antibodies specific to SARS-CoV-2 and the associated cytokines in colostrum from women who tested positive for the virus.

**Study Design:** Between March and September 2020 we obtained bilateral colostrum samples collected on spot cards within 48 hours of delivery from 15 new mothers who had previously tested positive for SARS-CoV-2. Five of these 15 COVID-19 positive women also provided bilateral liquid colostrum within 1-2 days of providing the spot card samples. Archived bilateral colostrum samples collected from 8 women during 2011-2013 were used as pre-COVID-19 controls. All samples were tested for reactivity to the Receptor Binding Domain (RBD) of the SARS-CoV-2 spike protein using an ELISA that measures SARS-CoV-2 RBD-specific IgA, IgG, and IgM, and for concentrations of 10 inflammatory cytokines (IFNγ, TNFα, IL-1β, IL-2, IL-4, IL-6, IL-8, IL-10, IL-12, IL-13) using a multiplex electrochemiluminescent sandwich assay.

**Results:** Bilateral colostrum samples from 73%, 73% and 33% of the 15 COVID-19 mothers exhibited IgA, IgG, and IgM reactivity to RBD respectively. Colostrum samples from two of the 8 pre-pandemic controls showed IgA and IgG reactivity to RBD. Additionally, COVID-19 mothers had significantly higher levels of 9 of the 10 inflammatory markers (all except IFNγ) as compared to the pre-COVID-19 controls. Comparable results were obtained with both the spot card-eluates and liquid samples.

**Conclusions:** A strong humoral immune response is present in the colostrum of women who were infected with SARS-CoV-2 before delivering. High levels of 9 inflammatory markers were also present in the colostrum. The evolution and duration of the antibody response, as well as dynamics of the cytokine response, remain to be determined. Our results also indicate that future large-scale studies can be conducted with milk easily collected on paper spot cards.

## INTRODUCTION

The Center for Disease Control and Prevention and the World Health Organization (WHO) recommend breast-feeding for mothers infected with SARS-CoV-2, as the benefits of mother’s milk are thought to outweigh potential risks of transmitting the virus to the infant^1,2^. A recent systematic review, reporting on 77 nursing mothers from 37 studies concluded that there was no convincing evidence of transmission of SARS-CoV-2 via breastmilk^3^. As of December 14^th^, 2020, the WHO reported over 71 million people infected by SARS-CoV-2 globally, and over 1.5 million deaths. As the number of pregnant and lactating SARS-CoV-2-infected women increases, there is a need to build on existing, yet limited, research on SARS-CoV-2-specific-antibodies and immune response in breast milk from infected women. Antibodies to SARS-CoV-2 and cytokines in breast milk are relevant to the health of nursed babies and mothers^1,2,4,5^. Multiple studies have reported an increase in inflammatory cytokines in the serum and bronchoalveolar lavage fluid of COVID-19-infected individuals^6–12^. However, there are no reports on the cytokine profiles in breast milk of women with COVID-19. Published literature suggests that the transfer of cytokines via breast milk can impact an infant’s immune system, conferring protection against various infectious diseases and allergies^13,14^. We^15,16^ and others^13,14,17^ have measured cytokines in breast milk and colostrum. Understanding cytokine profiles in breast milk of COVID-19-infected women is particularly relevant, as a preliminary study indicates that expression of ACE2 is elevated in the mammary epithelium during pregnancy, and through JAK-STAT pathways cytokines can influence *ACE2* promoter activation^18^. Antibody-mediated protection from SARS-CoV-2 in breast milk has clinical implications regarding the discussion about breast-feeding after infection. Prolonged antibody presence may ultimately influence the maternal decision to breast-feed and aid in the support a mother receives post-partum. The present study details findings regarding SARS-CoV-2-specific IgA, IgG, and IgM, and cytokine profiles in bilateral samples of colostrum collected within the first few days after parturition from 15 infected women and compares these results to colostrum obtained from pre-COVID-19 samples collected during 2011-2013.

## MATERIALS AND METHODS

### Recruitment of COVID-19-positive participants

Study participants were patients at UMass Memorial Medical Center (UMMC, Worcester, MA) and provided consent in accordance with an IRB-approved protocol (H00020140). Fifteen participants who tested positive, and one participant (P01) who tested negative for the SARS-CoV-2 RNA, provided bilateral colostrum on the day of, or the day after delivery. Participants hand-expressed colostrum from each breast onto spot cards (Whatman® FTA® card, Millipore Sigma, #WHAWB120205); which were left to dry at room temperature (RT).

Of the 16 participants who provided bilateral colostrum on spot cards, six participants also provided liquid bilateral colostrum within two days after providing the spot card samples. Participants hand-expressed or pumped colostrum equivalent to 5-10 mL from each breast into containers. The spot cards and liquid samples were stored at −80°C at UMMC until transferred to the laboratory at UMass Amherst at which point the spot cards were stored at RT and the liquid samples were stored at −20°C.

### Selection of pre-COVID-19 controls

We identified archived samples from eight women who donated liquid bilateral colostrum (1-3 days post-partum) during June 2011-May 2013. These colostrum samples were obtained following IRB-approved protocols.

### Processing bilateral colostrum samples on spot cards

Discs (6 mm diameter) prepared from spot cards were heat-treated for 30 minutes at 56°C to inactivate any virus and were then resuspended in 500 μL of Tris-buffered saline with 0.1% Tween 20^®^ (TBST) in a 24-well plate. The plate was incubated with gentle shaking overnight at 4°C after which the TBST-eluates were used for detection of anti-SARS-CoV-2-specific immunoglobulins. Extra eluates were stored at 4°C and used for the analysis of cytokines within 72 hours.

### Processing liquid bilateral colostrum from COVID-19 and pre-COVID-19 samples to obtain a whey fraction

Briefly, 500 μL of colostrum was centrifuged at 820g for 8 minutes. The whey fraction was transferred to a 2 mL centrifuge tube and heat-treated for 30 minutes at 56°C. Samples from the whey fraction were used for the detection of anti-SARS-CoV-2-specific immunoglobulins. Extra whey fractions were stored at 4°C and used for the analysis of cytokines within 72 hours.

### Enzyme-Linked Immunosorbent Assay (ELISA) for the detection of anti-SARS-CoV-2 IgA, IgG and IgM

A SARS-CoV-2 ELISA was developed and validated at UMass Amherst. The receptor binding domain (RBD) spike protein cloned into the pCAGGS expression vector was expressed in HEK293T cells (ATCC) using PEI (10:1 PEI:DNA ratio) and purified by gravity flow, as described in Stadlbauer *et al*^19^. Briefly, 96-well plates (Fisher Sci., #351172) were coated with the RBD spike protein at 1 μg/mL in 1X phosphate-buffered saline and incubated with gentle shaking overnight at 4°C, followed by blocking in 5% (w/v) dry skimmed milk in TBST with gentle shaking for 30 minutes at RT. Fifty microliters of sample were added and incubated with gentle shaking for 1 hour at RT. Wells were then washed with TBST and incubated with horseradish peroxidase (HRP)-conjugated goat anti-human-IgA, goat anti-human-IgG, or goat anti-human-IgM at 1 μg/mL (Jackson Laboratory). Plates were washed 3 times, incubated with 2,2’-Azinobis [3-ethylbenzothiazoline-6-sulfonic acid]-diammonium salt (ATBS; Sigma Aldrich, #A9941) diluted at 0.2 mg/mL in 0.1 M Sodium Acetate pH 4.5 at 37^0^C for 30 minutes. Known concentrations of anti-Spike-RBD-human (h) IgG1, -hIgM and -hIgA1 (InvivoGen, San Diego, CA, #C3022) were assayed in the ELISAs. The concentration of the highest standard was 1250 ng/mL; subsequent standards were prepared by 10-fold serial dilutions starting from 500 ng/mL to 0.05 ng/mL. After background subtraction, concentration curves for IgA, IgG and IgM were generated using a four-parametric logistic (4PL) curve with Excel’s Solver Add-In. Concentrations of unknown samples were calculated using the 4PL equation.

### Analysis of cytokines

Cytokines were measured in a multiplex assay (Mesoscale Discovery, Gaithersburg, MD) according to the manufacturer’s instructions using 10-plex human V-PLEX Proinflammatory Panel 1 plates. Each 96-well plate included an 8-point standard curve and assays for ten cytokines: IL-2, IL-4, IL-6, IL-8, IL-10, IL-12p70, IL-13, IL-1, IFN-*γ*, TNF-*α* (upper and lower limits of detection are in **Table S1**). Samples and standards were run in technical duplicates.

### Data Analysis

Welch’s t-test was used to assess differences in age and BMI between donations made during 2020 and during 2011-2013. Student’s t-test was used to compute differences between cytokines. *P*-values < 0.05 were considered statistically significant after Bonferroni correction. Multiple linear regression using the *lm*() function was performed in R (version 4.0.2) to assess correlations between levels of anti-RBD IgA, IgG and IgM and cytokines.

## RESULTS

### Participant Demographics

Demographic characteristics of the 24 women are summarized in **Table 1**, stratified by the period of colostrum donation: ‘2020’ (COVID-19) versus ‘2011-2013’ (Pre-COVID-19 controls). Groups did not differ significantly by age or BMI. Colostrum collected during 2011-2013 was exclusively from women who identified as White (one woman did not provide information on race). Women who provided colostrum in 2020 were more diverse: they self-identified as 31% Hispanic, 13% White Hispanic, 50% non-Hispanic White, and one woman identified as Asian American.

**Table 1.**
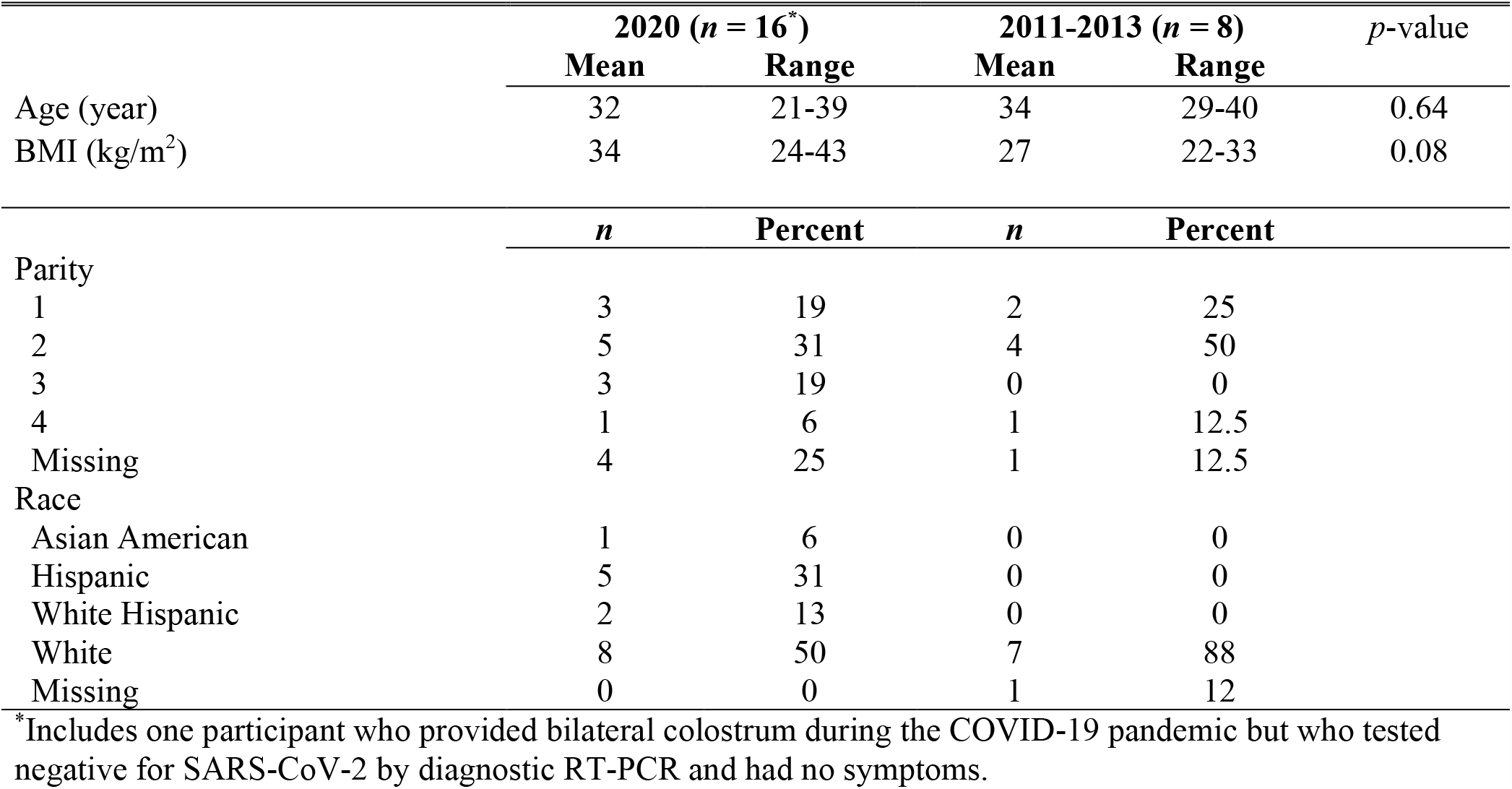
Demographics of participants who donated colostrum

|  | 2020 (*n* = 16<sup>*</sup>) |  | 2011-2013 (*n* = 8) |  | *p*-value |
| --- | --- | --- | --- | --- | --- |
|  | Mean | Range | Mean | Range |  |
| Age (year) | 32 | 21-39 | 34 | 29-40 | 0.64 |
| BMI (kg/m<sup>2</sup>) | 34 | 24-43 | 27 | 22-33 | 0.08 |
|  | *n* | Percent | *n* | Percent |  |
| Parity |  |  |  |  |  |
| 1 | 3 | 19 | 2 | 25 |  |
| 2 | 5 | 31 | 4 | 50 |  |
| 3 | 3 | 19 | 0 | 0 |  |
| 4 | 1 | 6 | 1 | 12.5 |  |
| Missing | 4 | 25 | 1 | 12.5 |  |
| Race |  |  |  |  |  |
| Asian American | 1 | 6 | 0 | 0 |  |
| Hispanic | 5 | 31 | 0 | 0 |  |
| White Hispanic | 2 | 13 | 0 | 0 |  |
| White | 8 | 50 | 7 | 88 |  |
| Missing | 0 | 0 | 1 | 12 |  |

\*Includes one participant who provided bilateral colostrum during the COVID-19 pandemic but who tested negative for SARS-CoV-2 by diagnostic RT-PCR and had no symptoms.

Eleven of the 15 participants tested positive for COVID-19 near the time of delivery (0-4 days (**Figure 1**). The 4 participants who did not test positive near the time of delivery (P13, P14, P15, and P16), had their most recent positive test 16 to 116 days before delivery (**Figure 1**). Six of the 15 COVID-19^pos^ participants reported no symptoms. All of the participants reporting no symptoms (P02, P04, P05, P08, P10 and P11) had their first positive test within 1-3 days of delivery. Onset of symptoms for the remaining 9 participants occurred between 27-144 days before delivery (**Figure 1**). Eight of the 9 participants reported that onset of symptoms occurred at the time of the first positive test; the ninth participant (P09) reported onset of symptoms 18 days before her first positive test.

**Figure 1.**
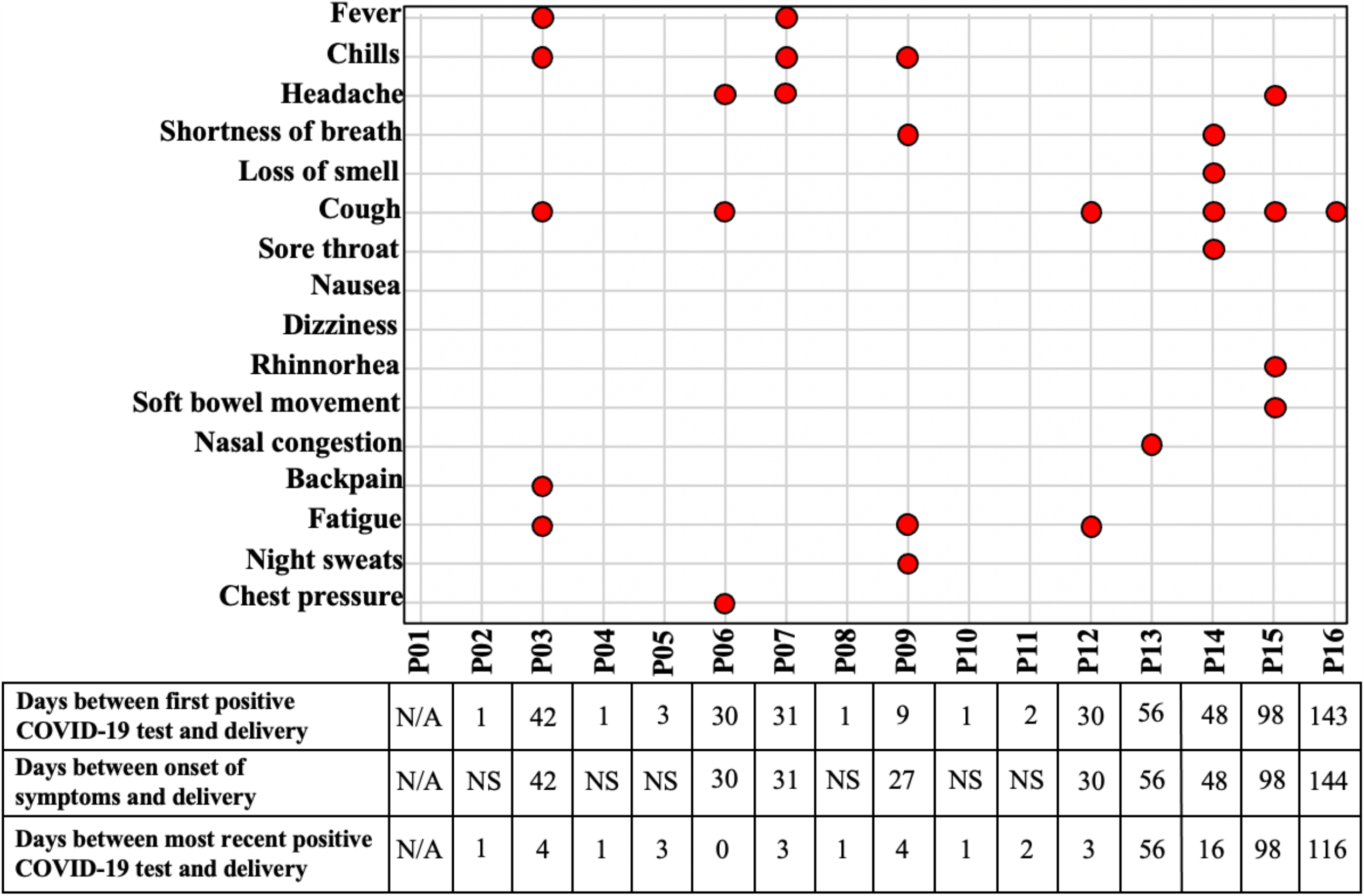
Overview of participants’ COVID-19 symptoms relative to their time of delivery. Participant P01 tested negative for SARS-CoV-2 (indicated as not applicable: N/A). Participants P02, P04, P05, P08, P10, and P11 reported no COVID-19-related symptoms (indicated as no symptoms: NS) despite positive PCR tests.

### Colostrum obtained from COVID-19^pos^ participants exhibited strong reactivity to anti-RBD IgA, IgG, and IgM

IgA, IgG, and IgM reactivities were measured in bilateral colostrum samples on spot cards. Samples were collected within 48 hours of delivery from 15 women who had positive SARS-CoV-2 (COVID-19^pos^) tests, and one woman (P01) who tested negative for SARS-CoV-2. Five of the 15 COVID-19^pos^ participants and P01 also provided liquid bilateral colostrum samples within 2 days after providing the spot card sample. Eight liquid bilateral colostrum samples donated as part of other studies during 2011-2013 served as pre-pandemic controls. All samples were tested in technical replicates. The low mean CVs of 3.21%, 3.9% and 4.4% for IgA, IgG and IgM assays respectively (**Figure S1**) demonstrate the high precision of the assay.

Positive cut-off values for each assay were set at twice the mean OD levels for secondary-only antibody reactivities (background). The binding reactivities of IgA, IgG and IgM were similar between colostrum obtained from left and right breasts (**Figure 2A, B, and C**). For spot cards, **Figures 2A & 2B** show that colostrum was reactive to the RBD spike protein in samples from 14 of 15 participants for IgA and IgG, and 6 of 15 for IgM. The colostrum from only one COVID-19^pos^ participant (P15), had no reactivity to the RBD spike protein. Of the 6 participants who provided both spot card and liquid colostrum samples (P01-P06), the overall reactivities appear similar, but with a few differences. Reactivities for IgA, IgG and IgM were higher in the first donation (spot card) than in the second donation (liquid) for P03 and P06, while this pattern was reversed for P04 (**Figure 2A**).

**Figure 2.**
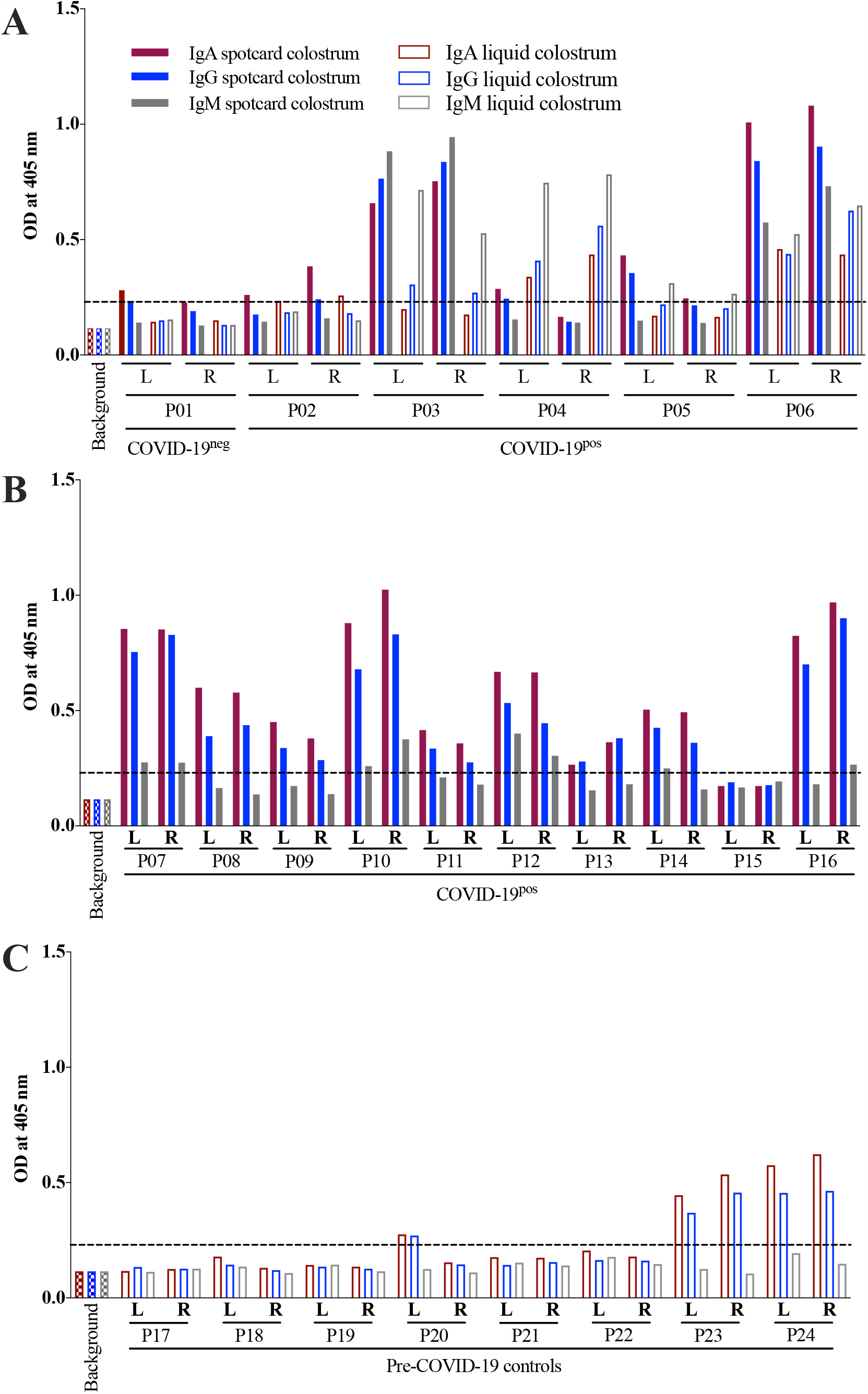
Distinct reactivities for IgA, IgG and IgM in colostrum from COVID-19-infected and pre-COVID-19 control participants. (A) Mean OD values for IgA, IgG and IgM in colostrum from six participants who provided both bilateral colostrum on spot cards (filled bars, donation 1) and bilateral liquid colostrum (open bars, donation 2). (B) Mean OD values for IgA, IgG and IgM in colostrum obtained from the remaining 10 participants who provided bilateral colostrum on spot cards only. (C) Mean OD values for IgA, IgG and IgM in bilateral liquid colostrum obtained from 8 pre-COVID-19 control participants. For A and B, all spot card samples were collected within 48 hours postpartum and all liquid colostrum was collected 1-2 days after the spot card collection. For C, all liquid samples were collected 1-3 days postpartum. Dotted lines indicate cut-off value set at twice the mean OD of secondary-only antibody reactivity across all plates. OD values for all samples are the means of technical duplicates.

There is a strong relationship between having experienced symptoms and time since first positive test. Six participants (P02, P04, P05, P08, P10, and P11) had their first positive diagnostic test within 1-3 days of delivery, and none reported any symptoms, whereas all of the participants whose first diagnostic test was >25 days before delivery reported symptoms (**Figure S2**). In contrast, neither the time since first positive test nor whether the participant experienced symptoms were related to the antibody reactivity to RBD spike protein. Among women who tested positive at the time of delivery, the highest reactivities occurred in P06 and P10, who had their first positive test 30 days (symptomatic) and 1 day (asymptomatic), respectively, before delivery (**Figure S2**). Among the 6 women who did not have a positive test at delivery (P03, P09, P13 and P15 tested negative at delivery; P14 and P16 were not re-tested at delivery), all of whom had symptoms, the highest reactivity to RBD occurred in P03 and P16, who had their first positive test 42 and 144 days, respectively before delivery, while P15 had no reactivity and her first positive test 98 days before delivery (**Figure S2**).

Bilateral colostrum from 2 of 8 pre-COVID-19 control participants (P23 and P24), exhibited reactivities for IgA and IgG (**Figure 2C**). Colostrum from the left breast of pre-COVID-19 control, P20, also exhibited reactivities for IgA and IgG, albeit low (**Figure 2C**). Because we did not have a dilution factor associated with antibody extraction from the spot card, the statistical comparison of antibody levels between the COVID-19^pos^ and pre-COVID-19 controls was restricted to liquid samples.

The median IgA, IgG and IgM concentrations were 22.25 ng/mL versus 12.02 ng/mL; 8.47 ng/mL versus 4.50 ng/mL; and 93.89 ng/mL versus 30.27 ng/mL in colostrum obtained in 2020 and during 2011-2013 respectively (**Figure S3**). The greatest difference in antibody levels between COVID-19 participants and pre-COVID-19 controls was for IgM (p<0.0001), which showed a 3-fold higher concentration in the COVID-19^pos^ samples.

### Elevated inflammatory markers in colostrum from COVID-19^pos^ participants

Cytokines were measured in bilateral colostrum from COVID-19^pos^ and pre-COVID-19 participants. Among the COVID-19^pos^ participants, most analytes were detected in the majority of samples, and this was true for cytokines measured in liquid milk as well as in spot card-eluates (**Table 2**). In contrast, among the pre-COVID-19 controls, only one analyte, IL-8, was detected in all samples (**Table 2**).

**Table 2.** Concentrations of cytokines in human colostrum

| Analytes | COVID-19 Participants: March & September, 2020<sup>1</sup> |  |  |  |  |  | Pre-COVID Controls: 2011-2013 |  |  |
| --- | --- | --- | --- | --- | --- | --- | --- | --- | --- |
|  | Spot Card Colostrum *n* = 30<sup>2</sup> |  |  | Liquid Colostrum *n* = 10<sup>3</sup> |  |  | Liquid Colostrum *n* = 16<sup>4</sup> |  |  |
|  | Mean | IQR | % Det<sup>5</sup> | Mean | IQR | % Det | Mean | IQR | % Det |
| IFNγ | 2.78 | 0.78-3.34 | 84.4 | 20.55 | 2.22-26.61 | 91.7 | 13.38 | 0.14-11.17 | 69 |
| TNFα | 2.11 | 0.67-2.98 | 100 | 9.32 | 2.56-11.66 | 100 | 0.53 | 0.06-0.54 | 75 |
| IL-1β | 2.20 | 0.25-1.67 | 84.4 | 6.16 | 2.47-6.61 | 100 | 0.18 | 0.05-0.20 | 81 |
| IL-2 | 1.49 | 0.48-1.91 | 100 | 3.38 | 0.44-5.67 | 91.7 | 0.19 | 0.05-0.21 | 69 |
| IL-4 | 0.24 | 0.07-0.32 | 93.8 | 0.76 | 0.04-1.19 | 75 | 0.02 | 0.01-0.02 | 50 |
| IL-6 | 0.72 | 0.13-0.83 | 84.4 | 53.44 | 4.96-88.75 | 100 | 1.66 | 0.11-1.08 | 81 |
| IL-8 | 498 | 92-758 | 100 | 1045 | 511-1512 | 100 | 96.93 | 20 - 57 | 100 |
| IL-10 | 0.49 | 0.16-0.68 | 96.7 | 2.67 | 0.28-3.71 | 100 | 0.11 | 0.02-0.09 | 69 |
| IL-12 | 0.86 | 0.36-0.96 | 96.7 | 2.69 | 0.21-4.09 | 75 | 0.16 | 0.04-0.08 | 50 |
| IL-13 | 5.81 | 2.79-7.87 | 100 | 16.47 | 5.38-22.44 | 100 | 2.17 | 0.89-2.22 | 81 |

Concentration is pg/mL for all analytes. If both technical replicates were above the lower limit of detection (LLOD), the final concentration was the mean of the two levels. If both technical replicates were below the LLOD, the final concentration was computed as LLOD divided by the square root of 2<sup>29</sup>
<sup>1</sup> = Bilateral colostrum provided by 15 COVID-19-positive participants.
<sup>2</sup> = Spot card samples from the left and right breasts of 15 women diagnosed with COVID-19 collected within 48 hours of delivery
<sup>3</sup> = Liquid samples from the left and right breasts of 5 women diagnosed with COVID-19 collected 1-2 days after collection of the spot card samples
<sup>4</sup> = Liquid samples from the left and right breasts of 8 women who donated colostrum during 2011-2013 collected 1-3 days after delivery
<sup>5</sup> = Percent of samples with detectable analytes

Cytokine concentrations were higher among COVID-19^pos^ participants as compared to pre-COVID-19 controls, and again this was the case for both liquid colostrum and spot card-eluates (except for IFN*γ* and IL-6, which were not higher in the spot card). However, the concentration of cytokines was higher in the liquid colostrum than in the spot card-eluates for all analytes (**Table 2**). Because we did not have a dilution factor associated with the spot card extraction, statistical comparison between COVID-19^pos^ and pre-COVID-19 samples was limited to the liquid colostrum.

Among liquid samples, significantly elevated concentrations for 9 of 10 cytokines were observed in the COVID-19^pos^ group (2020) (**Figure 3**). Only IFN*γ* was not significantly higher. Moreover, there is an indication that cytokine levels in bilateral colostrum obtained from symptomatic participants were higher (red-filled circles; **Figure 3**) compared to levels in asymptomatic participants. This distinction was particularly clear for IL-2, IL-4, IL-6, IL-10 and IL-12. Additionally, we explored the association between antibody response and cytokine levels and found that SARS-CoV-2-specific IgA, IgG and IgM were negatively correlated to IL-13 in spot card-eluates (**Table 3**). This pattern was not detected in liquid colostrum from pre-COVID-19 controls (**Table S2**).

**Table 3.**
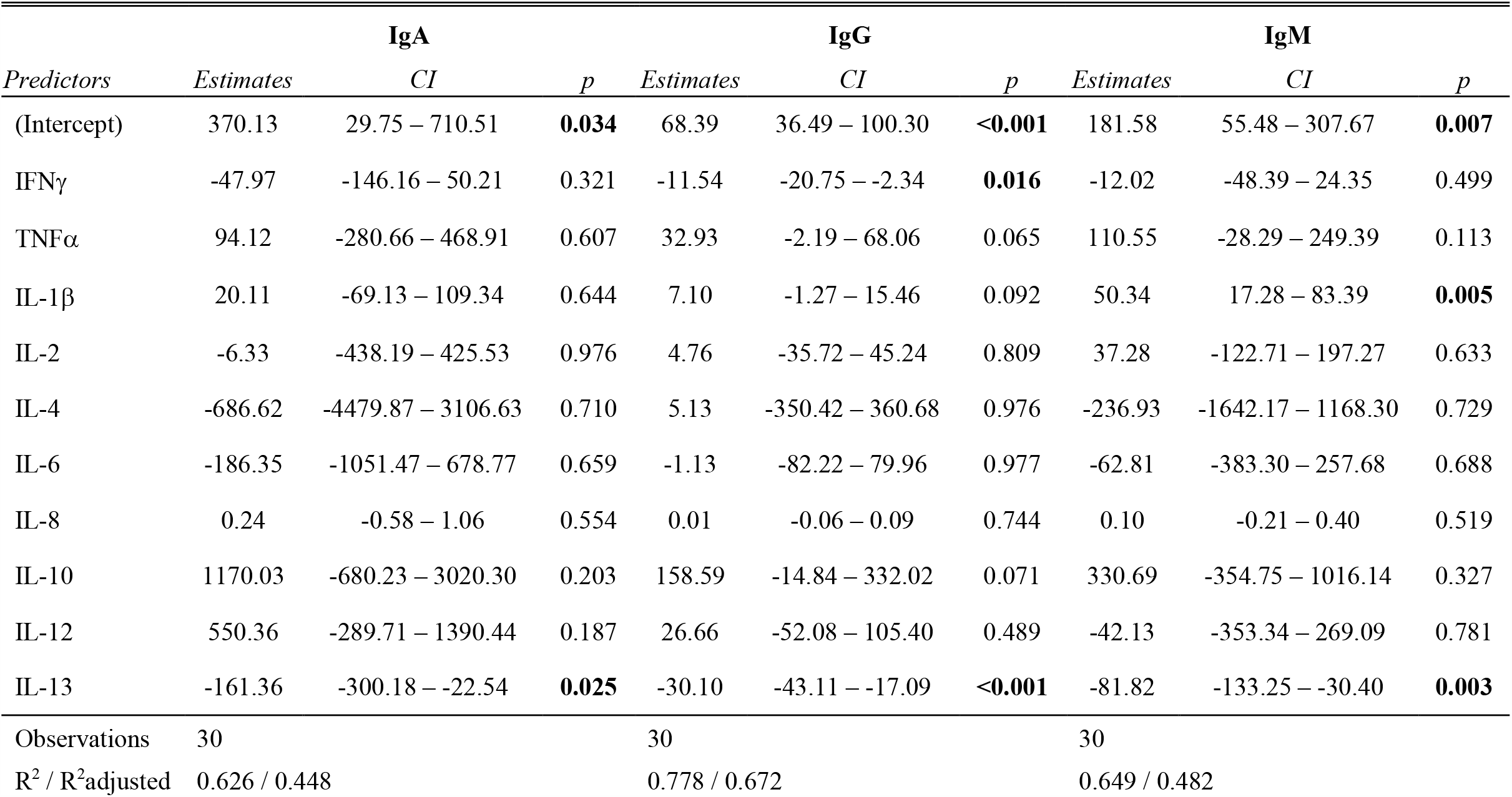
Correlation of IgA, IgG and IgM concentrations with cytokine concentration in bilateral colostrum eluted from spot cards obtained from 15 participants diagnosed with COVID-19

**Figure 3.**
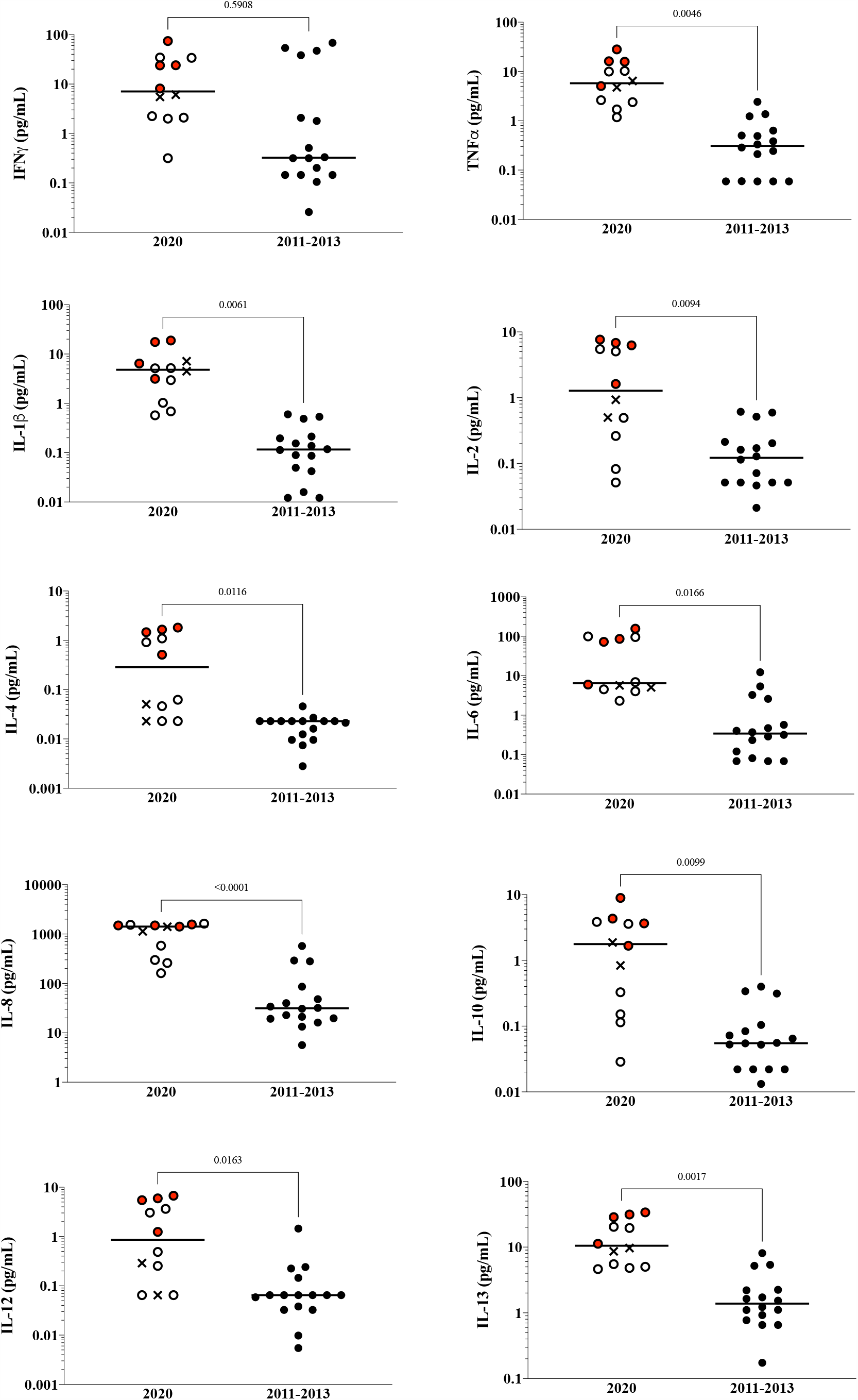
Elevated inflammatory markers in colostrum obtained from COVID-19 participants. Concentrations of cytokines in liquid colostrum from the five COVID-19 participants, the 2020 COVID-19-negative participant (**x**), and the pre-COVID-19 pandemic controls (●) were measured with MSD technology. The plots show the median concentration of each cytokine (middle line). Red circles 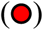 indicate bilateral colostrum provided by participants who exhibited COVID-19-related symptoms. Open circles (○) below median values for all analytes indicate bilateral colostrum provided by participants P02 and P04 who did not exhibit COVID-19-related symptoms despite having positive PCR tests. Open circles above median values for all analytes indicate bilateral colostrum provided by participant P05 who did not exhibit COVID-19-related symptoms but reported having influenza 3 months before delivery.

## DISCUSSION

### Principal findings

Our results provide a snapshot of the dynamic immune response in colostrum following SARS-CoV-2 infection, confirming recent findings on the presence of SARS-CoV-2-specific antibodies in milk from infected women^4,5^, and describing for the first time, the associated cytokine profile. Colostrum samples archived before the pandemic together with analysis of bilateral samples provide important controls for this study, and the similarities between results from the spot card and the liquid colostrum demonstrate the value of the spot card collection method.

Spot card colostrum from all but one of the 15 participants exposed to SARS-CoV-2 exhibited IgA, IgG and IgM reactivities to RBD. The similarity in reactivity levels between the two breasts provides confidence in the assay; comparable levels of immunoglobulins across breasts are expected^20^ except when there are local infections. The range of IgA, IgG and IgM reactivities to RBD is not easily explained by the time since onset of symptoms. The single COVID-19^pos^ participant whose colostrum had no antibody reactivity to RBD (P15) had onset of symptoms 98 days before delivery, while one of the participants with high levels of IgA and IgG reactivities to RBD (P16) had onset of symptoms 144 days before delivery (**Figure S2**).

### Results in the context of what is known

The presence of SARS-CoV-2-specific IgA and IgG in bilateral liquid colostrum of two pre-COVID-19 controls, and the left breast of a third control (P20), could indicate a prior infection that elicited a humoral response that cross-reacted with SARS-CoV-2-RBD. This would be consistent with findings from Pace *et al*, who demonstrated that levels of SARS-CoV-2-specific milk IgA and IgG correlated with IgA and IgG concentrations specific to the S-protein of 229E coronavirus^4^. Alternatively, Isho *et al* also observed elevated saliva IgA and IgG reactivities to SARS-CoV-2-RBD among their pre-COVID-19 controls, but did not attribute the response to a prior coronavirus infection^21^. We were concerned that the reactivity in the left breast of P20 could have been due to experimental error, however, a repeat analysis confirmed the IgA and IgG reactivities.

Interestingly, IFN*γ* is the only cytokine that was not elevated in the colostrum of COVID-19^pos^ participants. Many viruses, including SARS-CoV-2, have developed mechanisms to evade the antiviral type-1 interferon pathway, leading to its reduced expression and moderate to low expression of various interferon stimulated genes, including IFN*γ*^6,22–25^. While there are some conflicting reports, the lack of increased IFN*γ* expression associated with COVID-19 is consistent with our findings in colostrum and needs further investigation.

An inverse relationship between IL-10 and proinflammatory cytokines is normal and expected, as IL-10 is upregulated in T-cells in response to inflammation and acts to reduce inflammatory cytokines. However, in SARS-CoV-2 infections, there is evidence that cytokines are not secreted by T-cells, but by recruited monocytes and macrophages, causing IL-10 to remain elevated^26–28^. We show that both IL-10 and inflammatory cytokines are increased in colostrum. A time-course study with serial milk collections and analysis of both cytokines and immune cells would contribute greatly to our understanding of the inflammatory response to SARS-CoV-2 infections.

Spot cards provide an efficient means of collecting colostrum. All 16 participants consented to provide spot card followed by liquid colostrum. That the liquid sample was collected from only 6 participants is likely due to the smaller volume requested for the spot card, and greater ease for the staff, as the spot card can be left to dry at RT for several hours, while the liquid colostrum needs to be quickly frozen.

### Clinical Implications

The possible protection from SARS-CoV-2-specific antibodies detected in colostrum has clinical implications regarding the discussion about breast-feeding after exposure to the virus. SARS-CoV-2-specific immune response was detected in the colostrum of women who had their first positive test and symptoms more than four months before delivery, women who were symptomatic at delivery, as well as asymptomatic women who had a first positive test at delivery. The detection of SARS-CoV-2-specific antibodies in these women with diverse COVID-19 disease experiences provides objective data for the value of initiating breast-feeding despite SARS-CoV-2 infection.

### Research Implications

Given that the differences between the spot card and liquid samples for IgA, IgG and IgM reactivities to RBD are not all unidirectional, we can assume that differences do not simply reflect collection method. Indeed, the differences between spot card and liquid reactivities for P04 could demonstrate antibody evolution in colostrum: i.e., a switch from high IgA to high IgM (**Figure 2A**).

Cytokine concentrations are significantly higher in the liquid colostrum from COVID-19^pos^ as compared to pre-COVID-19 participants (**Figure 3**) suggesting a SARS-CoV-2-specific response. However, there is concern that cytokines could be degraded in archived samples. An argument against sample degradation is the high levels of IFN*γ* for two of the pre-COVID-19 controls.

Calculated concentrations of cytokines differed between the spot card-eluates and liquid colostrum among the five COVID-19^pos^ participants who provided both sample types, with the levels being generally higher in the liquid samples (**Figure S4**). Four participants show an increase in the liquid samples, and one participant shows a decrease. Differences could reflect rapid changes in levels over days. Use of the spot card with serial collections and direct comparisons between liquid and spot card samples are needed in future work.

## Conclusion

Our study is among the first to demonstrate the presence of SARS-CoV-2-specific antibodies in colostrum and describes for the first time elevated cytokines in colostrum from women exposed to SARS-CoV-2. The evolution and duration of the antibody response as well as dynamics of the cytokine response remain to be determined. Given the feasibility of the collection method, and the ability to detect antibodies and cytokines, our results indicate that future large-scale studies can be conducted with milk easily collected on paper spot cards.

## Data Availability

The authors confirm that the data supporting the findings of this study are available within the article [and/or] its supplementary materials.

## DECLARATIONS

### Ethics approval and consent to participate

Approved by IRBs at UMMC to HL (H00020140) and at UMass Amherst to KA (2075)

### Consent for publication

Informed consent was obtained from all patients

## Authors’ contributions

KA, HL, VN, and BP conceptualized the overall study design. KA, DA, VN, and BP, designed and optimized analysis methods. HL, EC, AK, KM, and TL assisted with collection of samples. VN performed all laboratory experiments and statistical analyses. KA, VN, and BP prepared the first draft of the manuscript. All authors read and edited earlier versions and approved the final manuscript.

## Acknowledgements

We are grateful to all participants who donated colostrum for this study.

