## Supplemental figures for "Humoral and cell-mediated response in colostrum after exposure to severe acute respiratory syndrome coronavirus 2"

Figure S1

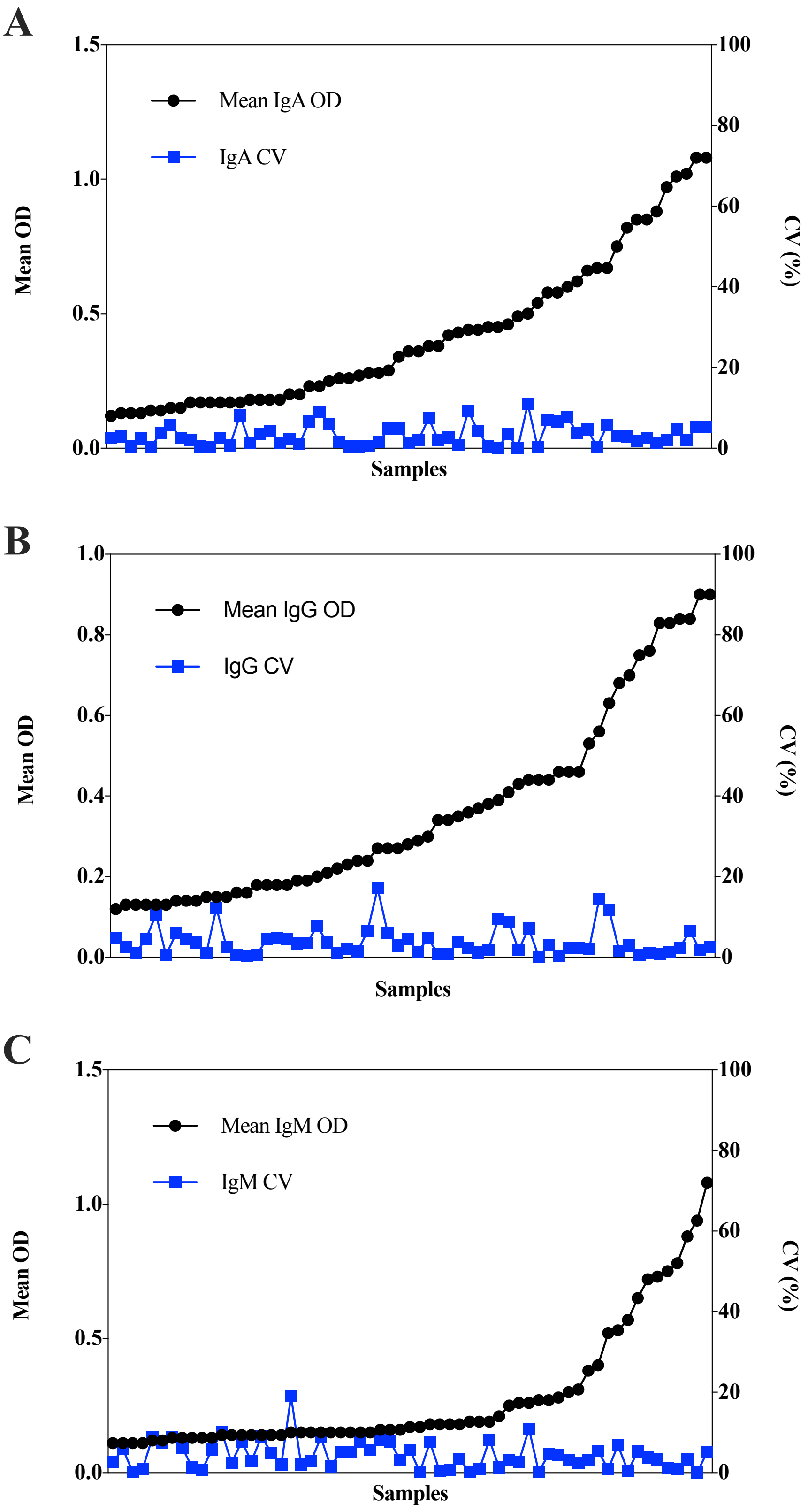

A
1.5
1.0
0.5
0.0
Mean OD
Mean IgA OD
IgA CV
Samples
100
80
60
40
20
0
CV (%)
B
1.0
0.8
0.6
0.4
0.2
0.0
Mean OD
Mean IgG OD
IgG CV
Samples
100
80
60
40
20
0
CV (%)
C
1.5
1.0
0.5
0.0
Mean OD
Mean IgM OD
IgM CV
Samples
100
80
60
40
20
0
CV (%)

**Figure S1. Performance evaluation of the SARS-CoV-2 specific IgA, IgG and IgM ELISA assay.** Reactivities to IgA (A), IgG (B) and IgM (C) (black circles) and their corresponding coefficient of variation (CV) (blue boxes) of the 60 colostrum samples (from the left and right breasts of 24 women). Each sample was tested in technical duplicate and the CV of duplicate OD values was calculated.

Figure S2

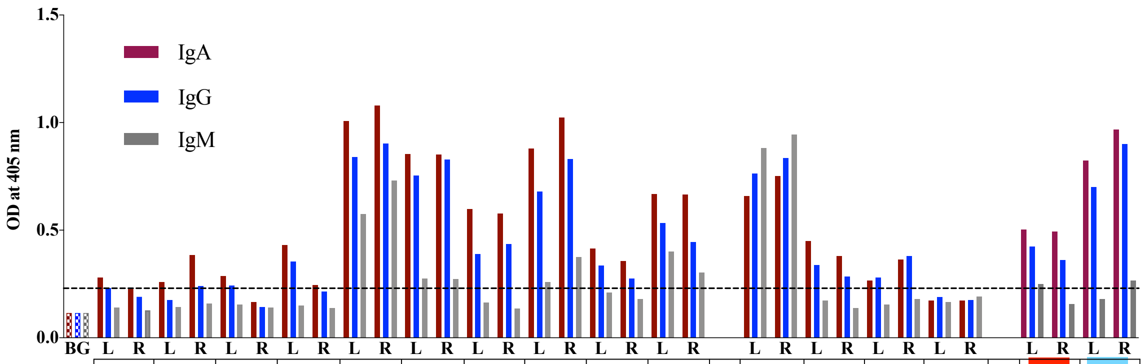


| | P01 | P02 | P04 | P05 | P06 | P07 | P08 | P10 | P11 | P12 | P03 | P09 | P13 | P15 | P14 | P16 |
|---|---|---|---|---|---|---|---|---|---|---|---|---|---|---|---|---|
| ***First +*** | N/A | 1 | 1 | 3 | 30 | 31 | 1 | 1 | 2 | 30 | 42 | 9 | 56 | 98 | 48 | 143 |
| ***Symptom onset*** | N/A | NS | NS | NS | 30 | 31 | NS | NS | NS | 30 | 42 | 27 | 56 | 98 | 48 | 144 |
| ***Most recent +*** | N/A | 1 | 1 | 3 | 0 | 3 | 1 | 1 | 2 | 3 | 4 | 4 | 56 | 98 | 16 | 116 |
| | **Positive for COVID-19 at delivery** | | | | | | | | | | **Negative for COVID19 at delivery** | | | | | |

***First +*** Days between first positive COVID-19 test and delivery

***Symptom onset*** Days between onset of symptoms and delivery

***Most recent +*** Days between most recent positive COVID-19 test and delivery

Not re-tested at delivery

Tested negative 101 days prior to delivery and was not re-tested at delivery

P01 is a COVID-19 negative 2020 participant

NS - No reported COVID-19 symptoms

**Figure S2. Overview of participants' first positive COVID-19 test, symptoms onset, most recent positive test and delivery.** Bars indicate mean OD values for IgA, IgG and IgM in colostrum from 16 participants who provided bilateral colostrum on spot cards. Participants were stratified based on whether they tested positive, negative or not determined (P14 and P16) at delivery. Participants P06, P07 and P12 tested positive 0-3 days prior delivery and experienced symptoms during their first positive test. Participants P03, P13 and P15 tested positive 4-98 days before delivery, but tested negative at delivery, and experienced symptoms during their first positive test. Participant P09 tested positive 4 days before delivery, but tested negative at delivery, and experienced symptoms 18 days prior to her first positive test.

Figure S3

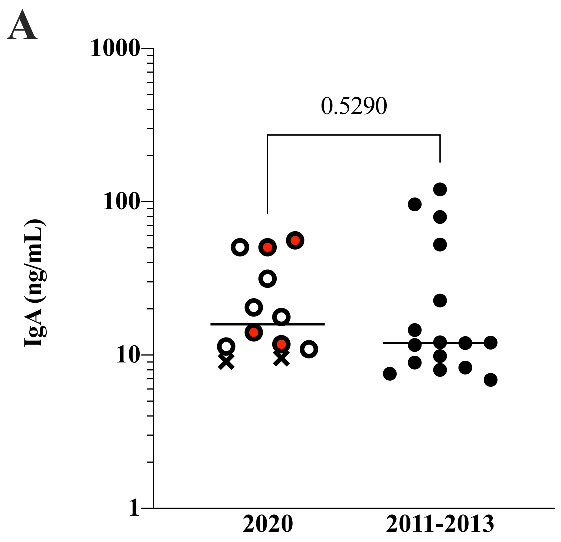

A
1000
100
10
1
IgA (ng/mL)
0.5290
2020
2011-2013


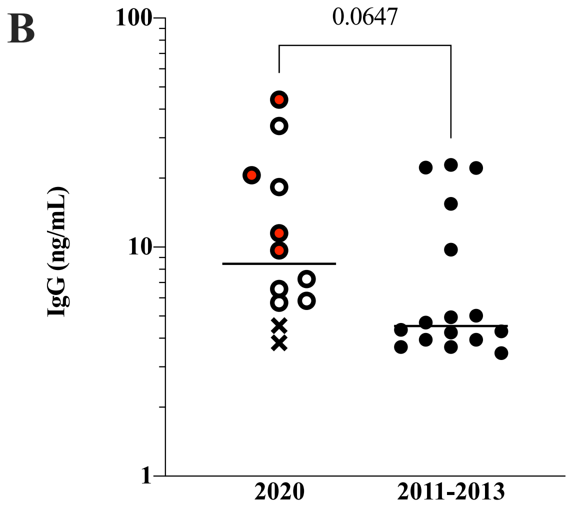

B
100
10
1
IgG (ng/mL)
0.0647
2020
2011-2013


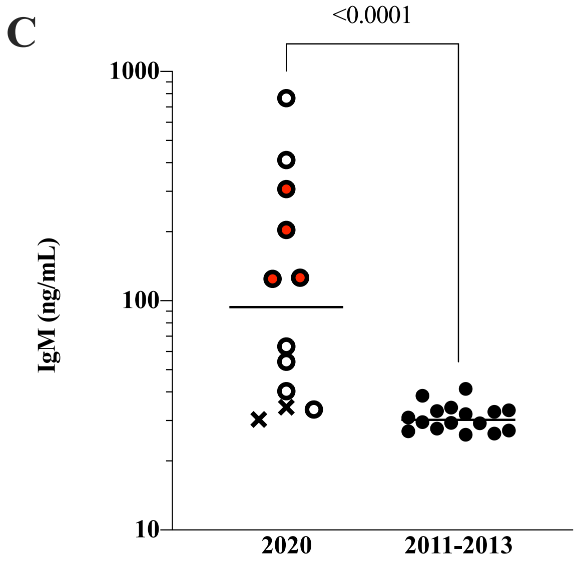

C
1000
100
10
IgM (ng/mL)
<0.0001
2020
2011-2013

**Figure S3. Elevated IgA, IgG and IgM in COVID-19 participants**. Concentrations of IgA (A), IgG (B) and IgM (C) were calculated from ODs obtained for liquid colostrum samples using a four-parametric logistic curve generated with Excel's Solver Add-In (see Methods). Participants who tested positive for COVID-19 (○,●) were compared to the pre-COVID-19 controls (●). The single 2020 COVID-19-negative participant (x) is included in the figure but was not included in the statistical analyses. Red circles (●) indicate bilateral liquid colostrum provided by participants who exhibited COVID-19-related symptoms.

Figure S4

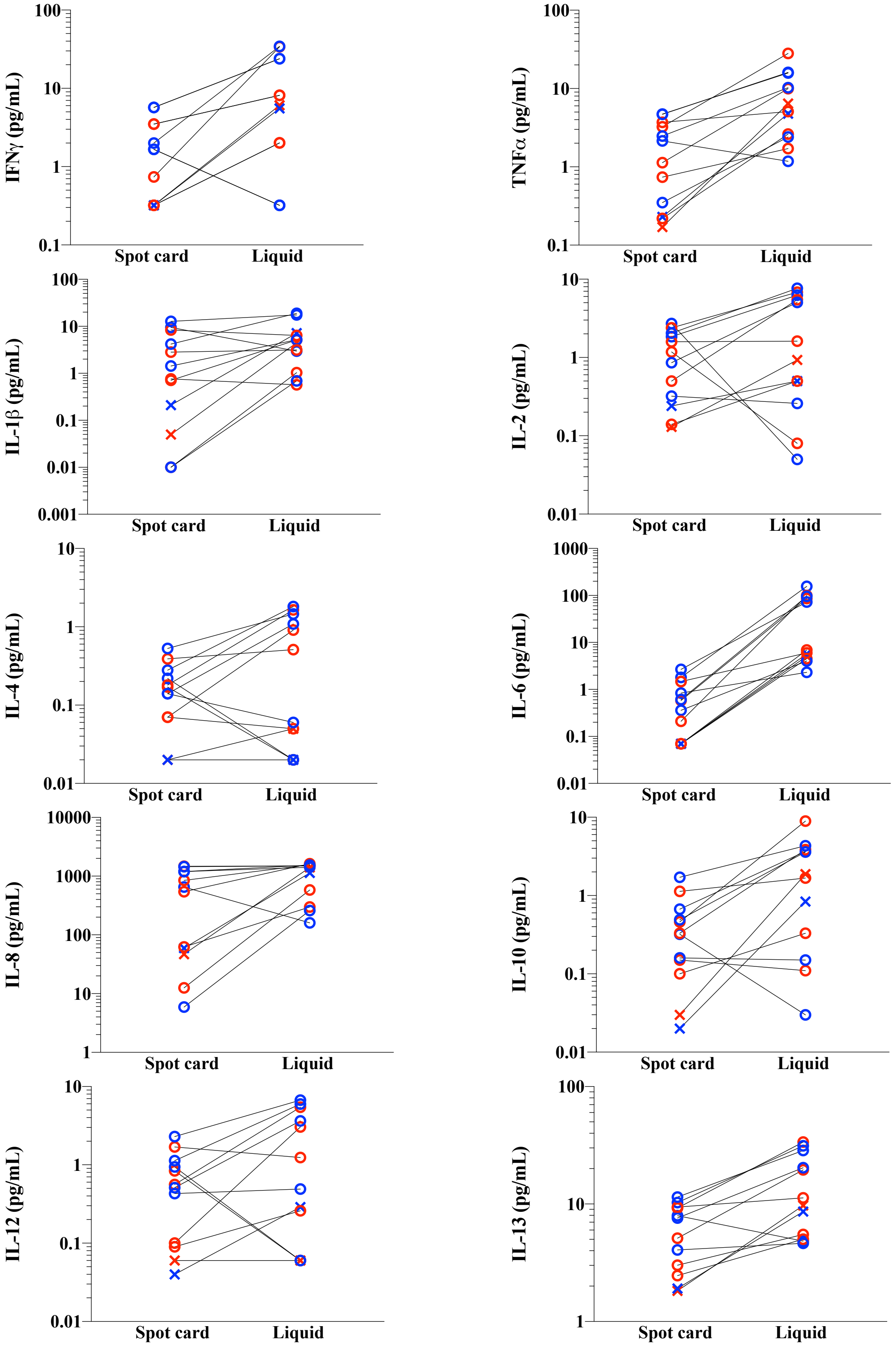

IFNγ (pg/mL)
TNFα (pg/mL)
IL-1β (pg/mL)
IL-2 (pg/mL)
IL-4 (pg/mL)
IL-6 (pg/mL)
IL-8 (pg/mL)
IL-10 (pg/mL)
IL-12 (pg/mL)
IL-13 (pg/mL)
Spot card
Liquid

**Figure S4. Cytokine concentrations in participants who provided both spot card and subsequent liquid colostrum (P01 – P06).** Blue circles indicate concentrations in colostrum from the right breast and red circles indicate concentrations in colostrum from the left breast. The symbol **x** indicates concentration in COVID-19 negative participant (blue: right breast, red: left breast).
