## Supplemental tables for "Humoral and cell-mediated response in colostrum after exposure to severe acute respiratory syndrome coronavirus 2"

**Table S1.** Lower and Upper limits of detection for the 10 analytes tested in the MSD assay

| | LLOD (pg/mL) | | ULOD (pg/mL) |
|---|---|---|---|
| **Analytes** | **Plate 1** | **Plate 2** | **Plates 1 and 2** |
| IFNγ | 0.45 | 0.20 | 1500 |
| TNFα | 0.08 | 0.04 | 367 |
| IL-1β | 0.02 | 0.02 | 610 |
| IL-2 | 0.07 | 0.03 | 1360 |
| IL-4 | 0.03 | 0.02 | 211 |
| IL-6 | 0.09 | 0.05 | 690 |
| IL-8 | 0.05 | 0.03 | 612 |
| IL-10 | 0.03 | 0.02 | 356 |
| IL-12 | 0.09 | 0.05 | 464 |
| IL-13 | 0.93 | 0.25 | 509 |

**Table S2**. Correlation of IgA, IgG and IgM concentrations with cytokine concentration in bilateral liquid colostrum obtained from 8 pre-COVID-19 control women.

| | **IgA** | | | **IgG** | | | **IgM** | | |
|---|---|---|---|---|---|---|---|---|---|
| *Predictors* | *Estimates* | *CI* | *p* | *Estimates* | *CI* | *p* | *Estimates* | *CI* | *p* |
| (Intercept) | 2.69 | -37.41 – 42.79 | 0.870 | 3.71 | -5.86 – 13.29 | 0.365 | 30.28 | 22.12 – 38.43 | **<0.001** |
| IFNγ | -0.26 | -1.20 – 0.69 | 0.516 | -0.10 | -0.33 – 0.12 | 0.295 | 0.00 | -0.19 – 0.19 | 0.968 |
| TNFα | -65.88 | -243.50 – 111.74 | 0.384 | -24.79 | -67.21 – 17.63 | 0.193 | -7.37 | -43.49 – 28.76 | 0.623 |
| IL-1β | -3.46 | -430.60 – 423.68 | 0.984 | 18.51 | -83.50 – 120.53 | 0.660 | 50.51 | -36.36 – 137.38 | 0.195 |
| IL-2 | 97.86 | -500.90 – 696.61 | 0.692 | 47.69 | -95.31 – 190.69 | 0.430 | 11.83 | -109.94 – 133.60 | 0.813 |
| IL-4 | 222.85 | -1419.39 – 1865.10 | 0.741 | 67.13 | -325.08 – 459.35 | 0.678 | -103.08 | -437.07 – 230.90 | 0.464 |
| IL-6 | 0.20 | -27.03 – 27.43 | 0.986 | 1.72 | -4.78 – 8.22 | 0.527 | 0.98 | -4.56 – 6.52 | 0.668 |
| IL-8 | 0.04 | -0.29 – 0.37 | 0.777 | -0.01 | -0.08 – 0.07 | 0.866 | -0.00 | -0.07 – 0.06 | 0.883 |
| IL-10 | 128.41 | -1102.21 – 1359.02 | 0.799 | -27.94 | -321.85 – 265.96 | 0.817 | -57.17 | -307.44 – 193.10 | 0.583 |
| IL-12 | 40.68 | 5.37 – 76.00 | **0.031** | 10.11 | 1.67 – 18.54 | **0.027** | -2.87 | -10.05 – 4.31 | 0.352 |
| IL-13 | 8.70 | -23.88 – 41.28 | 0.523 | 2.12 | -5.66 – 9.90 | 0.515 | 0.33 | -6.30 – 6.95 | 0.904 |
| TNFα | -65.88 | -243.50 – 111.74 | 0.384 | -24.79 | -67.21 – 17.63 | 0.193 | -7.37 | -43.49 – 28.76 | 0.623 |
| Observations | 16 | | | 16 | | | 16 | | |
| $R^2$ / $R^2$adjusted | 0.931 / 0.793 | | | 0.906 / 0.718 | | | 0.796 / 0.389 | | |
